# Clinical and bacterial determinants of unfavorable tuberculosis treatment outcomes: an observational study in Georgia

**DOI:** 10.1101/2025.01.20.25320828

**Authors:** Galo A. Goig, Chloé Loiseau, Nino Maghradze, Kakha Mchedlishvili, Teona Avaliani, Ana Tsutsunava, Daniela Brites, Sevda Kalkan, Sonia Borrell, Rusudan Aspindzelashvili, Zaza Avaliani, Maia Kipiani, Nestani Tukvadze, Levan Jugheli, Sebastien Gagneux

## Abstract

Tuberculosis (TB) remains a major public health concern. Improving TB control programmes and treatment success rates requires a deeper understanding of the factors that determine disease presentation and patient treatment outcomes. While the demographic and clinical factors influencing treatment outcomes are well documented, the role of bacterial genetics remains limited. In this study, we analyzed the *Mycobacterium tuberculosis* complex (MTBC) genomes and the associated clinical data from 4,536 TB patients in the country of Georgia covering a period of 13 years. Multivariable modelling confirmed the role of known demographic and clinical factors such as sex, age, body mass index (BMI) and comorbidities in determining treatment outcomes, as well as the efficacy of novel TB treatments containing bedaquiline. In addition, we found that some bacterial variables, including the MTBC lineage, the specific mutations conferring resistance to rifampicin and fluoroquinolones, as well as a high bacterial burden were associated with unfavorable outcomes. GWAS analyses revealed no genetic mutations in the bacteria other than known drug resistance-conferring mutations to be associated with treatment outcomes. However, we found that mutations in the bacterial gene *sufD* were linked to cavitary disease. Additionally, we observed that mutations in *sufD,* mutations conferring resistance to rifampicin and fitness compensatory mutations were associated with the bacterial burden within patients. We conclude that both patient and bacterial factors determine disease presentation and clinical outcomes in TB.

## Introduction

Despite tuberculosis (TB) being preventable and treatable, it remains one of the top ten leading causes of human mortality worldwide, and the leading cause of death from a single infectious agent[1,2]. For susceptible TB, treatment success rates are high, yet globally, more than 12% of TB patients fail treatment[2]. The situation is considerably worse for drug-resistant forms of TB, with success rates averaging only 68% for multidrug-resistant TB (MDR-TB), and 44% for extensively drug-resistant TB (XDR-TB)[2,3]. Recently, WHO endorsed the use of regimens based on the novel and repurposed drugs bedaquiline, pretomanid and linezolid, with or without moxifloxacin (BpaL(M))[4], that have improved treatment outcomes for MDR-TB[5]. However, despite these BPaL(M) regimens being introduced only recently, resistance to the new drugs is already emerging and spreading between patients[6]. Therefore, in order to preserve the life-span of these new regimens and at the same time strengthen TB control programmes, there is a need to develop tailored interventions that can improve treatment success rates. For this we need to better understand the factors that influence treatment outcomes of TB patients across populations and epidemiological settings.

The clinical and demographic factors that influence treatment outcomes are generally well understood. Specifically, comorbidities such as HIV, diabetes or alcohol abuse, patient characteristics such as age, sex or body mass index (BMI), and social factors such as unemployment, imprisonment, or homelessness are well documented (refer to Peetluk *et al.* for a systematic review on modeling studies[7]). In contrast, factors related to the *Mycobacterium tuberculosis* Complex (MTBC), which is the main causative agent of TB, are less understood. A key bacterial factor is antibiotic resistance. It is well established that resistance to TB drugs leads to worse treatment outcomes[2]. However, drug resistance is not a binary trait, but a spectrum that ranges from full susceptibility to varying degrees of resistance. For example, it has been shown that small variations in the minimum inhibitory concentration (MIC) to isoniazid and rifampicin in pretreatment MTBC patient isolates were associated with patient relapse[8]. The level of drug resistance itself is influenced by the specific resistance mutations conferring resistance to the particular drug[9], as well as by the bacterial genetic background, in which the mutation occurs[10–12]. Therefore, genetic analyses of the infecting MTBC strains may provide opportunities to optimize treatments and improve treatment success rates. For example, several studies have shown that mutations associated with a high-level of resistance to fluoroquinolones were also associated with poor patient treatment outcomes [13–15]. However, our understanding of how different mutations conferring resistance to different TB drugs affect treatment success is still very limited. This is partly because most resistance is caused by only a few frequent mutations, while many other rare mutations often lack the necessary sample size for meaningful analyses[13–15]. Additionally, the association between specific resistance mutations and patient outcomes have mostly been studied without taking into consideration other key clinical and demographic factors. Therefore, we still lack a clear understanding of how the different resistance-conferring mutations may influence treatment outcomes in TB.

Apart from drug resistance-conferring mutations, other bacterial factors could also play a role in determining different clinical manifestations of TB and patient treatment outcomes. For example, “modern Beijing” strains have been associated with worse treatment outcomes[16], while strains belonging to MTBC Lineage 1, appear to be more prone to cause disseminated TB and bone disease[17,18]. However, only few studies have attempted to find novel genetic determinants of patient treatment outcomes [11,19–22], and most of these have been limited by small sample sizes. Moreover, few studies have analyzed the effect of bacterial genetics while controlling for relevant clinical and demographic factors. As a consequence, our current understanding of the overall clinical relevance of bacterial genetics in determining treatment outcomes and disease progression in TB remains limited.

In this study we analyzed 4,536 MTB genomes isolated from patients diagnosed with TB in Georgia over a period of 13 years. By integrating in our analysis comprehensive clinical and demographic patient data, we aimed to identify bacterial and host factors that are relevant to patient treatment outcomes and disease manifestation.

## Results

### Description of the study cohort

We analyzed MTBC clinical isolates from culture-confirmed TB patients diagnosed in the country of Georgia between 1st October 2010 and 31st December 2023. This included all MDR-TB patients for which an MTBC isolate was available, and all patients with drug-susceptible TB diagnosed between 2014 and 2016. In total, we analyzed the MTBC genomes and associated patient data from 4,536 TB patients. Of these, 4,368 (96%) genomes passed our quality criteria for downstream analysis. Out of the 4,368 TB patients with a high-quality MTBC genome, 3,579 (81.9%) had known treatment outcomes, 389 of which (10.9%) were unfavorable (defined as death, treatment failure or relapse). The overall median patient age was 39 years (interquartile range = 29-52y) and 3,144 patients (72.0%) were male. HIV-coinfection was determined negative for 3,544 patients (81%), positive for 145 patients (3%) and was unknown for 679 patients (16%). Based on our genomic analysis, 2,375 MTBC isolates (54.4%) were susceptible to all anti-TB drugs, 19 (0.4%) were mono/polyresistant, 94 (2.2%) were isoniazid mono-resistant, 53 (1.2%) were rifampicin mono-resistant, 1,338 (30.6%) were MDR, 466 (10.7%) were pre-extensively drug-resistant (MDR plus resistance to fluoroquinolones) and 23 (0.5%) were considered XDR (pre-XDR plus additional resistance to bedaquiline, delamanid or linezolid). Of the patients with drug-resistant TB, 1,579 (79.2%) were treated with fluoroquinolones, and 647 (32.5%) were treated with regimens containing bedaquiline, delamanid and/or linezolid. A complete description of the study population and the relationship of each of the 66 variables considered in this study is provided in the Supplementary Material. The Supplementary Material is publicly available in a GitLab repository, where we provide supplementary methods, results, the complete dataset, and a detailed step-by-step description of all analyses conducted in R markdown documents.

Link to GitLab repository: https://git.scicore.unibas.ch/TBRU/georgia_tb_tx_outcomes

### Description of the MTBC isolates and drug resistance-conferring mutations

Most MTBC isolates belonged to Lineage 2 (L2) (n=2,176; 49.8%) and L4 (n=2,098, 48.0%). These proportions changed among drug-resistant cases, in which L2 clearly dominated (n=1,546, 79.9%; Fisher test p-value<0.001). Most L2 strains belonged to L2.2.1 Central Asia (n=1,370, 63.0%; 35.3% among drug-resistant cases, Fisher test p-value<0.001) and to the L2.2.1 “W148” clade, which has been linked with MDR-TB outbreaks in Eastern Europe and Central Asia L2.2.1 W148[23] (n=707, 32.5%; 32.2% among drug-resistant cases; Fisher test p-value<0.001). The majority of L4 strains belonged to sublineages L4.8 (n=728; 16.7%), L4.3.3 (n=598, 13.7%), L4.2.1 (n=363; 8.3%), and L4.1.2 (n=276, 6.3%). In 78 isolates (1.8%), we detected different sublineages coexisting, and hence these cases were classified as mixed infections. Among isoniazid-resistant strains (n=1,915), nearly all strains carried the mutation KatG Ser315Thr (n=1,803; 94.2%). Among rifampicin-resistant strains (n=1,879), the most common rifampicin resistance-conferring mutation was RpoB Ser450Leu (n=1,323, 70.4%). More than half of rifampicin-resistant strains (n=1,017, 54.1%) also carried mutations compensating for the fitness cost of rifampicin resistance[24], and nearly all of these strains carried the RpoB Ser450Leu mutation (n=1,006, 98.9%). Among a total of 505 fluoroquinolone-resistant strains identified, the most common resistance mutations were GyrA Asp94Gly (n=181, 35.8%) and GyrA Ala90Val (n=165, 32.7%).

### Clinical and bacterial factors associated with TB treatment outcomes

To identify factors associated with treatment outcomes we analyzed all patients with available treatment outcome data (n=3,579; Supplementary Material). Briefly, we modeled the data by selecting relevant predictors based on least absolute shrinkage and selection operator (LASSO) regularization and variable importance according to a random forest, and then building a multivariable logistic regression model. Out of the 66 variables initially considered, 14 were included in the final model, which we refer to as the “base model”. The model performance assessed by 10-fold cross-validation showed an area under the receiving operator characteristic curve (AU-ROC) of 0.81 and an area under the precision-recall curve (AU-PRC) of 0.44 (0.11 for a random classifier). These metrics show that despite the dataset’s class imbalance (11% in the “unfavorable outcome” class), the model effectively captures a substantial part of the signal explaining treatment outcomes. In the base model, previous treatment failure, defaulting treatment, or relapse, being unemployed, male sex, older age, lower body mass index (BMI), presenting with dyspnea, hiv-coinfection, being treated for other diseases, any form of drug resistance, and shorter time-to-culture positivity (TTP) were associated with unfavorable outcomes, while being treated with regimens containing bedaquiline, delamanid and/or linezolid was strongly associated with favorable outcomes (Table 1). In general, cases with missing data for employment, drug abuse, dyspnea and hiv status were associated with unfavorable outcomes.

**Table 1.** Variables associated with unfavorable treatment outcomes in a multivariable logistic regression. We refer to this as the “base” model. BMI (body mass index); TTP (time-to-culture positivity; Bdq (bedaquiline); Dlm (delamanid); Lzd (linezolid).

| Variable | aOR<sup>†</sup> | 95% CI<sup>†</sup> | p-value |
| --- | --- | --- | --- |
| Patient age | 1.05 | 1.04, 1.06 | **<0.001** |
| TTP | 0.96 | 0.93, 0.98 | **0.001** |
| BMI | 0.92 | 0.88, 0.97 | **0.002** |
| Diagnosis date | 1.04 | 0.98, 1.10 | 0.2 |
| Case definition: previous treatment failure | 4.58 | 2.13, 9.68 | **<0.001** |
| Case definition: other | 1.45 | 0.90, 2.27 | 0.11 |
| Case definition: relapse | 1.82 | 1.28, 2.55 | **<0.001** |
| Case definition: treatment after defaulting | 4.73 | 3.08, 7.22 | **<0.001** |
| HH ratio | 0.95 | 0.76, 1.10 | 0.6 |
| Mono/Polyresistant | 5.18 | 1.51, 16.9 | **0.007** |
| INH Monoresistant | 3.56 | 1.49, 7.75 | **0.002** |
| RIF Monoresistant | 4.56 | 1.26, 14.1 | **0.012** |
| MDR | 9.75 | 5.65, 17.0 | **<0.001** |
| pre-XDR | 26.3 | 14.4, 48.9 | **<0.001** |
| XDR | 24.8 | 6.10, 108 | **<0.001** |
| Male | 1.68 | 1.24, 2.30 | **0.001** |
| Employed: missing | 3.41 | 1.80, 6.59 | **<0.001** |
| Employed: yes | 2.22 | 1.38, 3.76 | **0.002** |
| HIV-coinfection: missing | 1.49 | 1.06, 2.06 | **0.018** |
| HIV-coinfection: yes | 1.68 | 0.92, 2.97 | 0.082 |
| Drug abuse: missing | 2.94 | 1.90, 4.52 | **<0.001** |
| Drug abuse: yes | 0.64 | 0.19, 1.80 | 0.4 |
| Treatment for other diseases: missing | 0.89 | 0.48, 1.58 | 0.7 |
| Treatment for other diseases: yes | 1.78 | 1.16, 2.74 | **0.008** |
| Dyspnea: missing data | 2.59 | 1.25, 5.27 | **0.009** |
| Dyspnea: yes | 2.35 | 1.51, 3.62 | **<0.001** |
| Regimen containing Bdq/Dlm/Lzd | 0.23 | 0.14, 0.38 | **<0.001** |

### The association of MTBC lineage with TB treatment outcomes

We tested whether MTBC lineage was associated with treatment outcomes by adding lineage as a predictor to the base model (Supplementary Material). Since in Georgia most MTBC strains belonged to L2 and L4 (Figure 1), this predictor was divided into four levels: L2, L4, mixed infections, and uncommon lineages. In this model analyzing the full cohort, lineage was not a relevant predictor. However, because a previous meta-analysis found that Beijing strains (L2.2.1) were associated with poor treatment outcomes only among drug-susceptible cases[16], and because most L2 strains in Georgia belong to L2.2.1, we sought to replicate these results in our study. In support of these previous results, we found that L2 showed higher odds of unfavorable outcomes as compared to L4 among drug-susceptible cases (L2 adjusted odds ratio (aOR)=1.55; 95% confidence interval (CI)=1.05-2.27; p-value=0.026), but not among drug-resistant cases (aOR=0.87; CI=0.55-1.39; p-value=0.5; Supplementary Material). Additionally, we tested whether different MTBC sublineages may be associated with treatment outcomes but found no evidence for this (Supplementary Material).

**Figure 1.**
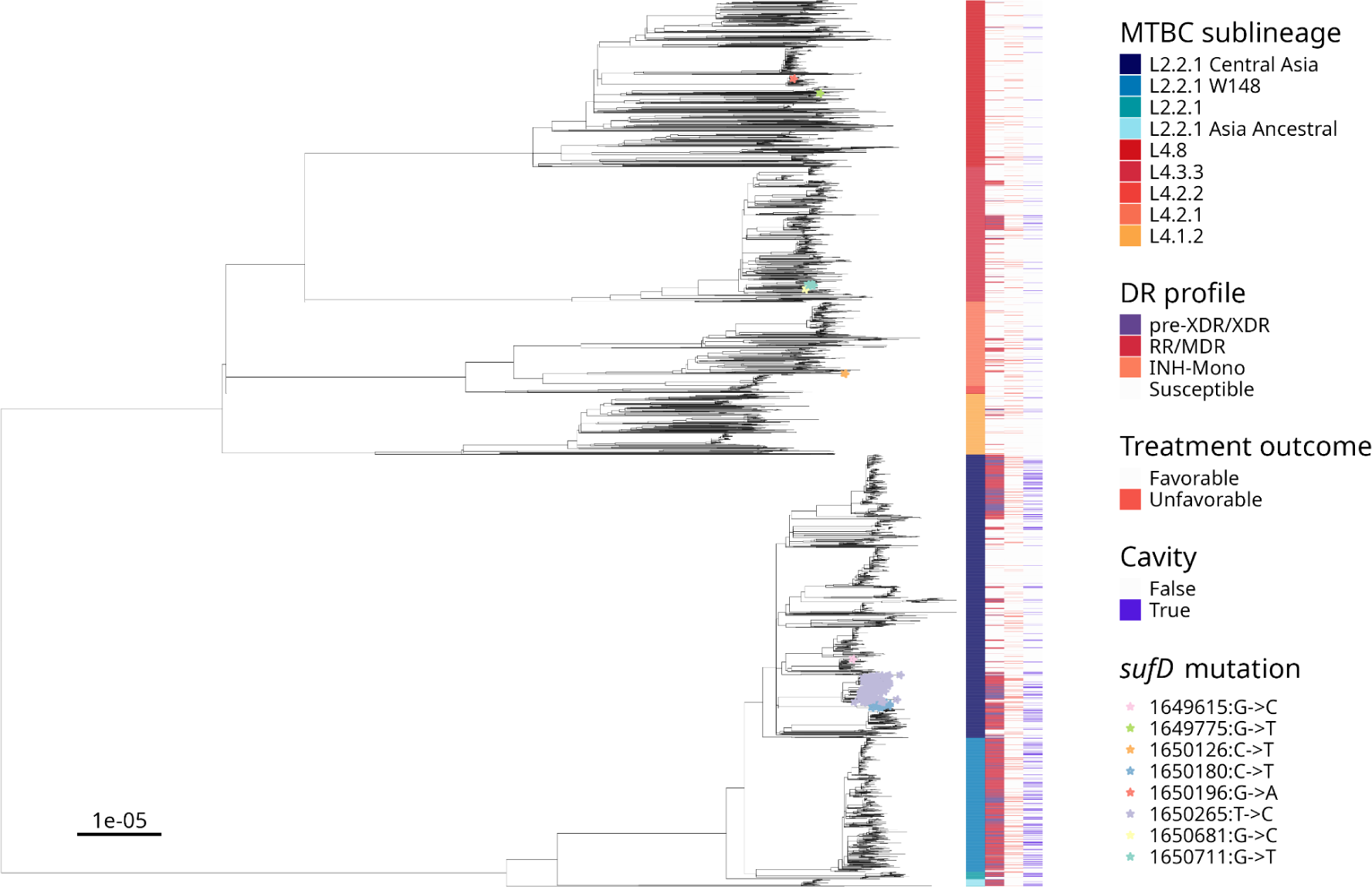
Phylogeny of 3,399 MTBC isolates from TB patients with favorable or unfavorable outcomes. The tree scale indicates substitutions per site. Mixed infections with different sublineages and minority sublineages are excluded for clarity. In the figure, the susceptible category of the drug resistance (DR) profile includes strains resistant to drugs other than isoniazid and rifampicin. Isolates carrying different non-synonymous mutations in *Rv1462* (*sufD*) are indicated with a coloured star. Notably, 57 strains carrying *sufD* mutations are excluded from the figure as their patient outcomes were neither favorable nor unfavorable.

### The association of mixed infections with TB treatment outcomes

Using the same analysis, we observed that mixed infections were not associated with unfavorable outcomes (Supplementary Material). However, in our initial models mixed infections were defined only based on the coexistence of different sublineages; a definition that does not consider infections caused by different MTBC genotypes belonging to the same sublineage. To address this, we used the ratio of unfixed to fixed SNPs as a measure of genetic diversity, which increases in isolates with multiple genotypes (Supplementary Material). Yet, even when measured by this ratio, the genetic diversity of the isolates was not associated with unfavorable outcomes in our cohort when assessed by univariate (t-test p-value=0.8) and multivariate analyses (aOR=0.95; CI=0.76-1.10; p-value=0.6).

#### The effect of specific drug resistance-conferring mutations on TB treatment outcomes

Using the base model, we further assessed whether specific mutations conferring resistance to different TB drugs could be differentially associated with unfavorable treatment outcomes. One problem with this analysis was that assessing the effect of many different drug resistance-conferring mutations led to a high degree of sub-stratification and to unstable models with unreliable estimates (Supplementary Material). To circumvent this, we built an alternative model, in which each individual mutation was substituted by the estimated MIC linked to this mutation based on a recent study from the CryPTIC consortium[9]. In this analysis, we excluded susceptible cases to avoid a naive modeling of the MIC as a near-binary predictor (MIC>0 yes/no). In this model, increasing MICs to fluoroquinolones (aOR=1.36 CI=1.22-1.50) and to rifampicin (aOR=1.28 CI=1.20-1.36) were associated with unfavorable treatment outcomes. Note that while isoniazid resistance was identified as a relevant predictor of unfavorable treatment outcomes (Supplementary Material), we were unable to assess the effect of different isoniazid-resistance mutations given that nearly all isoniazid resistance in this cohort is caused by the same mutation (i.e. KatG Ser315Thr). Similarly, the effect of MIC values to other relevant drugs such as bedaquiline or delamanid could not be assessed due to the low number observations.

Additionally, we tested whether heteroresistance, defined as the coexistence of both susceptible and drug-resistant strains within a patient, may be linked with treatment outcomes. Note that the analysis specifically assessed the effect of heteroresistance (presence or absence), independent of the effect of drug resistance itself. In univariate analysis, heteroresistance to any drug was associated with unfavorable outcomes (Fisher test p-value=0.018). However in multivariable analysis, this association was lost (Supplementary Material). We then tested whether heteroresistance to individual drugs may be associated with treatment outcomes. In univariate analysis, heteroresistance to fluoroquinolones was associated with unfavorable outcomes (Fisher exact test p-value=3.5e-5), and isoniazid heteroresistance showed a potential association with favorable outcomes (Fisher exact test p-value=0.07). However, in multivariable analyses only the potential association of isoniazid heteroresistance with favorable outcomes remained supported (aOR=0.23; CI=0.04-0.89; p-value=0.065; Supplementary Material).

#### GWAS of variants associated with TB treatment outcomes and disease manifestation

To determine whether genetic mutations other than those causing drug resistance could be associated with variable treatment outcomes or disease manifestation such as cavitation, dissemination and infiltration, we carried out multiple GWAS using pyseer[25]. These analyses were adjusted for both the clinical covariates and the MTBC population structure using the phylogeny of all strains. Specifically, we performed GWAS for individual genetic variants, as well as gene burden tests. After correcting for multiple testing, we did not find any MTBC genetic variant associated with treatment outcomes (n=3,579 patients with treatment outcome data; Supplementary Material). Note that drug resistance mutations do not show as associated with outcomes, as the GWAS was adjusted for all the relevant covariates, including drug resistance.

To look for bacterial genetic variants associated with disease manifestation, we analyzed the full cohort (n=4,145, excluding mixed infections and sublineages with less than 30 observations to avoid over-stratification). After adjusting for multiple testing, we did not find any association with disseminated disease or infiltration. However, we found that non-synonymous mutations in the gene *Rv1462* (*sufD*) were associated with lower odds of cavitary disease (aOR=0.9; CI=0.86-0.95; p-value=1.86E-05). Inspection of the MTBC phylogeny showed that most strains carrying non-synonymous mutations in *sufD* belonged to two closely related drug-resistant subclades of L2.2.1 “Central Asia”, each one carrying a different mutation: SufD Val247Ala (n=153; 78.4%) and SufD Arg219Trp (n=29; 14.9%), respectively (Figure 1). The effect of these mutations in reducing the odds of cavitary disease may be linked to drug resistance, as cavitary disease appeared to be less common among drug-resistant strains carrying *sufD* mutations (Figure 1).

### Mutations in *sufD*, rifampicin resistance and compensatory evolution are linked to bacterial burden

To further explore our findings, we hypothesized that if strains carrying *sufD* mutations are less likely to cause cavitary disease, patients carrying strains with *sufD* mutations might present with a lower bacterial burden. To test this, we used TTP as a proxy for bacterial burden; TTP refers to the amount of days required for the MGIT culture system to detect growth of MTBC bacteria, and is directly linked to the bacterial load in the diagnostic sputum sample. We therefore built a model to find factors influencing TTP (Table 2; n=2,105 isolates with TTP data available; Supplementary Material). We found that psychiatric disorder, previous contact with a drug-resistant case, being treated with bedaquiline delamanid and/or linezolid, being treated with fluoroquinolones, and mutations associated with higher RIF MICs were associated with a longer TTP (i.e. lower bacillary load), whereas older age, male sex, receiving treatment for other diseases, presenting infiltration, dissemination, and cough, as well as being infected with MTBC strains carrying compensatory mutations, were associated with a shorter TTP (i.e. higher bacillary load) (Table 2).

**Table 2:** Factors associated with time to culture positivity among all samples with TTP data (n=2,105). *Bdq (bedaquiline); Dlm (delamanid); Lzd (linezolid)*.

| Variable | Estimate<br>(days) | 95% CI<sup>†</sup><br>(days) | p-value |
| --- | --- | --- | --- |
| Patient age | -0.02 | -0.04, 0.00 | **0.018** |
| Male | -1.0 | -1.7, -0.43 | **<0.001** |
| HIV-coinfection: missing | -0.35 | -1.2, 0.51 | 0.4 |
| HIV-coinfection: positive | 1.5 | -0.19, 3.3 | 0.082 |
| Treated with Bdq/Dlm/Lzd | 1.0 | 0.16, 1.9 | **0.021** |
| Treated with fluoroquinolones | 1.4 | 0.17, 2.7 | **0.027** |
| Compensated | -1.6 | -2.5, -0.66 | **<0.001** |
| Treatment for other diseases: missing | 0.35 | -1.0, 1.8 | 0.6 |
| Treatment for other diseases: yes | -1.2 | -2.3, -0.04 | **0.043** |
| Drug abuse: missing | -0.04 | -1.2, 1.1 | >0.9 |
| Drug abuse: yes | 1.7 | -0.51, 4.0 | 0.13 |
| Genotypic RIF MIC | 0.26 | 0.08, 0.44 | **0.004** |
| Genotypic EMB MIC | 0.33 | -0.05, 0.72 | 0.091 |
| Cough | -1.7 | -3.1, -0.40 | **0.011** |
| Dissemination | -1.5 | -2.4, -0.53 | **0.002** |
| Infiltration | -1.1 | -2.1, -0.15 | **0.024** |
| Psychiatric disorder | 4.1 | -0.12, 8.4 | 0.057 |
| Previous contact with DR case | 1.9 | 0.31, 3.4 | **0.019** |
| Case definition: previous treatment failure | -2.1 | -4.6, 0.51 | 0.12 |
| Case definition: other | 0.97 | -0.30, 2.2 | 0.14 |
| Case definition: relapse | 0.61 | -0.23, 1.5 | 0.2 |
| Case definition: treatment after defaulting | 0.03 | -1.3, 1.3 | >0.9 |
| *sufD* mutations | 1.3 | -0.13, 2.7 | 0.076 |

Being infected with strains carrying non-synonymous mutations in *sufD* also seemed potentially associated with TTP (beta=1.3 days, CI=-0.13-2.7; p-value=0.076). Since we observed in the phylogeny (Figure 1) that the effect of *sufD* may be linked to drug resistance, we repeated the regression analysis only among drug-resistant cases with available TTP data (n=905). We found that, in addition to the predictors described above, non-synonymous mutations in *sufD* (beta=1.7 days, CI=0.22-3.2; p-value=0.024) and MTBC L2 (beta=1.5 days, CI=0.50-2.6; p-value=0.004) were associated with a longer TTP (Supplementary Material).

## Discussion

We carried out an analysis of MTBC genomes and associated clinical data from patients diagnosed with TB in Georgia over a 13 year period to assess which patient and bacterial factors were relevant in influencing clinical outcomes and disease manifestation. Our results highlight the multifactorial nature of TB disease and confirm the role of known clinical and demographic variables in influencing patient outcomes. In addition, our findings support a role for bacterial factors. Specifically, we found that the MTBC lineage as well as the specific drug resistance-conferring mutations associated with varying levels of resistance influence treatment outcomes. Additionally, we show that various bacterial features, including lineage, compensatory mutations, and mutations in *sufD* are linked to the bacterial burden within patients and their likelihood of presenting with cavitary disease, probably through effects on within-host bacterial fitness.

Our work shows that patient factors such as older age, male sex, lower BMI, previous history of TB disease, unemployment, drug abuse, higher bacterial loads and drug resistance determine treatment outcomes in Georgia. These results align with existing evidence (reviewed in [7]). Moreover, this strong impact of patient factors implies that studies aiming at identifying bacterial factors influencing treatment outcomes need to control for relevant patient covariates. An important aspect of the cohort analyzed here is that a third of all patients with drug resistance were treated with novel drugs (i.e bedaquiline, delamanid and/or linezolid). Encouragingly, our results showed a substantial decrease in the odds of unfavorable outcomes when these drugs were used (aOR=0.23; CI=0.14-0.38), providing additional evidence of the efficacy of these novel regimens against drug-resistant TB[6,26].

Among the various bacterial factors that we assessed, enhanced levels of drug resistance were found to increase the odds of unfavorable outcomes (Table 1). Similarly, a higher bacterial burden, as indicated by a shorter TTP, was associated with unfavorable outcomes, indicating that patients harboring a higher bacterial burden, independently of drug resistance, are more likely to fail their treatment. These findings are also in line with previous studies[3,7,27,28].

We found MTBC L2 to be associated with unfavorable outcomes, but only among drug susceptible cases, which is in line with results from a previous systematic review and meta-analysis[16]. We see two potential explanations for these results, which are not mutually exclusive. First, it is possible that L2 strains are overall more virulent and associated with poor outcomes, and that the reason this association is not detected among drug-resistant isolates is because drug resistance itself has a much stronger effect on treatment outcomes than the MTBC lineage. Second, these results may suggest that L2 strains better survive first-line treatment, or are more likely to acquire drug resistance-conferring mutations, therefore leading to treatment failure and relapse in susceptible cases. These results add to a growing body of evidence showing that certain MTBC lineages are associated with traits relevant for disease outcomes, such as differences in susceptibility to TB drugs[11,12], or disease manifestation[17,18], and that therefore the phylogenetic diversity of the MTBC can be relevant for patient outcomes.

Contrary to previous work[29–31], we did not find mixed infections to be associated with unfavorable outcomes, both when considering the co-occurrences of defined sublinages or the genetic microdiversity in the sample. Similarly, we did not find an association between heteroresistance and unfavorable treatment outcomes. Previous studies have reported contradictory results regarding the association of mixed infections and heteroresistance with treatment outcomes[30–37]. While mixed infections and heteroresistance can lead to misdiagnosis and contribute to unfavorable patient outcomes[38–40], our results suggest that, as compared to other more relevant factors, they likely play a limited role on a broader scale.

One important finding in our study are the individual effects on treatment outcomes of specific mutations linked to different levels of resistance to fluoroquinolones and rifampicin. Although several studies have looked at this previously[13–15,41], they have been limited in sample size and did often not adjust for relevant patient covariates. In our analysis, instead of stratifying the resistance-conferring mutations into different categories, we modelled the estimated effect of these mutations on the respective MIC. When adjusting for clinical and demographic factors, we found that mutations conferring higher MICs for rifampicin and fluoroquinolones were associated with unfavorable outcomes. Our results support the fact that the efficacy of TB drugs does not depend on the bacterial resistance on a strict binary basis, but on the actual level of resistance. As high-dose isoniazid regimens are already endorsed to treat infections with isoniazid-resistant strains carrying low-level resistance mutations in *inhA*[42], and considering that regimens including high-dose isoniazid or rifampicin have been shown to be generally safe[42–44], our results support the rationale of optimizing treatments for drug-resistant TB with existing drugs.

Beyond drug resistance, we did not find any association between specific MTBC genetic variants and treatment outcomes. These results are not surprising given our stringent analysis, adjusting for both the population structure and relevant patient covariates. Our analytical approach meant that the effect size of any variant identified in the GWAS would be comparable to that of these patient covariates. These results are in agreement with benchmarks of GWAS methods using MTBC genomic data[45], but contrast with previous published studies performing GWAS, many of which have reported associations between MTBC genetic variants and treatment outcomes[11,19–22]. However, these associations were often identified using univariate analyses or observed in only a small number of strains. Moreover, none of these associations have so far been replicated in any independent patient cohort.

In contrast to the lack of bacterial associations with patient treatment outcomes, we found several bacterial features linked to key disease characteristics, including bacterial burden and the presence of lung cavities. Mutations linked to higher MICs to rifampicin were associated with longer TTP, reflecting a smaller bacterial burden. This observation aligns with previous research showing that drug resistance often imposes a replicative fitness cost on the bacteria[24,46]. Conversely, compensatory mutations mitigating the fitness cost of rifampicin resistance were associated with higher bacterial burdens, supporting a previous study in which we reported an association between the presence of compensatory mutations and positive smear microscopy[47]. These results suggest that the specific drug resistance mutations, as well as their epistatic interactions with other mutations, can have measurable effects on patients that go beyond the effect of drug resistance itself.

Finally, our study revealed non-synonymous mutations in *sufD* to be associated with lower bacterial burdens and lower odds of cavitary disease, particularly among drug-resistant cases. These results are compatible with existing evidence from different bacterial species, showing that *sufD* is an essential gene within the SUF system, which plays a critical role in redox metabolism and iron uptake[48,49]. Deficiencies in this system lead to increased susceptibility to oxidative stress and reduced bacterial survival[50,51]. Existing literature also shows an interplay between iron metabolism (and *sufD* in particular) with antibiotic resistance. For example, iron availability was shown to influence resistance to isoniazid, rifampicin or fluoroquinolones in different bacteria, including MTBC (reviewed in [52]). Experimental work has shown *sufD* to be upregulated after rifampicin exposure in *M. smegmatis*[53,54], as well as in *E. coli* strains carrying *rpoB* mutations[55]. In an experimental evolution setting, two out of 13 *E. coli* strains evolving in media with antibiotics acquired non-synonymous mutations in *sufD*[56]. These results are particularly interesting in the light of a recent study by Gorityala *et al.*, which aimed at identifying novel drug candidates against the MTBC[57]. After a comprehensive analysis, the authors pointed to *sufD* as an ideal candidate given its essential role in iron metabolism, but also to the availability of compounds binding this protein with a predicted low toxicity for humans. Taken together, these observations suggest that mutations in *sufD* may mediate a trade-off between virulence and antibiotic resistance, highlighting *sufD* as a potential drug target that could prove useful for treating DR-TB.

Our study has several limitations. One limitation is the approach used to estimate the effect of individual drug resistance-conferring mutations on treatment outcomes. The MIC values were assigned to each mutation based on a recent study that analyzed phenotypic MIC data for 13 different drugs in 15,000 MTBC strains to find genotype to phenotype associations[9]. However, MTBC strains carrying the same drug resistance-conferring mutation can show differences in MIC for the same drug[13], likely due to epistatic interactions between the drug resistance mutation and the strain genetic background[10]. However, the estimated MICs were determined based on a large sample size[9], and the MTBC samples analyzed here also included nearly 2,000 drug-resistant strains, which together should minimize potential biases in this analysis. Another limitation was the small number of observations for some variables, which translated into estimates with a wide confidence interval. This was particularly the case for mono/poly-resistance (n=17, aOR=5.18; CI=1.51-16.9) and XDR (n=14; 24.8; CI=6.19-108). Of note, pre-XDR also showed a wide confidence interval despite a high number of observations (n=334; aOR=26.3; CI=14.4-48.9). This uncertainty is probably related to a combination of over-stratification in the model and a strong effect size for the pre-XDR category. As pre-XDR is mostly defined based on fluoroquinolone resistance, this uncertainty when using drug resistance categories highlights the advantage of using alternative approaches such as modeling the MIC as we did in our study, where the model including fluoroquinolone MICs showed a narrower confidence interval for unfavorable outcomes (aOR=1.36; 95% CI=1.22-1.50).

In conclusion, our combined analysis of patient and bacterial variables from a large cohort of TB patients in Georgia over a 13-year period highlighted key demographic, clinical and bacterial determinants of TB treatment outcomes. In addition to confirming known patient factors, we showed that the MTBC lineage and specific drug resistance-conferring mutations were associated with treatment outcomes, and that additional bacterial variables such as the presence of compensatory mutations and mutations in *sufD* can influence disease presentation. Our results underscore the need to consider both patient and bacterial factors when optimizing TB treatment regimens.

## Methods

### Study cohort and samples analyzed

Between January 2011 and December 2023, *M. tuberculosis* isolates from all bacteriologically confirmed MDR-TB cases were collected and cultured at the National Center for Tuberculosis and Lung Disease in Tbilisi (NCTLD), Georgia. Additionally, isolates from all *M. tuberculosis* cases (including susceptible cases), were collected between the years 2014-2016. MGIT isolates were subcultured in 7H10 solid media plates. DNA was extracted from three loops of bacterial cells using a phenol-chloroform extraction and subsequently sent for whole-genome sequencing on Illumina NovaSeq 6000 and Illumina HiSeq 2500 platforms at the genomic core facility of the University of Basel and the Department of Biosystems Science and Engineering at ETHZ in Basel, Switzerland. Pseudo-anonymized patient-related data were routinely collected at NCTLD. Treatment outcomes were defined based on WHO guidelines[58]. In this study we specifically aimed to evaluate potential associations of bacterial factors with treatment outcomes; consequently, patients whose treatment outcome was classified as “lost to follow-up” were excluded. Treatment outcomes were defined as favorable for cases that were cured or that completed treatment, and unfavorable for those who showed treatment failure or died. Case definitions were also determined based on the WHO guidelines[59].

### Ethics approval

The National Ethics Council under the Ministry of Health of Georgia and the Local Ethics committee of the National Centre for Tuberculosis and Lung Disease in Tbilisi (Georgia), and the Ethics Commission of North- and Central Switzerland granted ethical approval for this study. The ethics committees waived the need for individual patient consent since only limited and anonymized clinical data were used.

### Whole-genome sequencing analysis

FASTQ files were processed with Trimmomatic[60] v0.39 to remove sequencing adaptors, trim low quality reads and keep reads longer than 20bp. For paired-end data, reads were merged using SeqPrep[61] v1.3.1 with an overlap size of 15bp. The resulting reads (both merged and unmerged) were mapped to the inferred chromosome of the *Mycobacterium tuberculosis* complex ancestor[62] using BWA mem[63] v0.7.17. Duplicates were identified and removed with Picard[64] v2.26.2. Sequencing reads were taxonomically classified using Kraken[65], and non-MTBC mappings were discarded as described previously[66]. Variants were called using the microbial mode of GATK[67] Mutect2 v4.2.4.1, and annotated and their effect predicted using SnpEff v4.1[68]. Additionally, we excluded from analysis supplementary and secondary alignments[69], and genomic positions in repetitive regions such as PE, PPE, and PGRS genes or phages[70]. Samples with an average sequencing depth lower than 20X or with more than 1% of contaminating reads from non-tuberculous mycobacteria as classified by Kraken were excluded from downstream analysis.

To identify drug-resistance conferring mutations, we compared all mutations detected in each genome with at least 10% allele frequency with the second edition of the catalog released by the WHO[71]. We only considered DR-conferring variants with a final confidence grading of “Assoc w Resistance” or “Assoc w Resistance – Interim”. We considered isolates to be pre-XDR if they showed resistance to rifampicin, isoniazid and any fluoroquinolone. We considered isolates to be XDR if they showed a pre-XDR genotype, and additionally resistance to one of the following: bedaquiline, delamanid, or linezolid. An estimated MIC for each drug was assigned based on the results of a previous study[9]. Whenever more than one drug resistance-conferring mutation was observed, we assigned the higher MIC. MTBC lineages were identified based on SNPs as described in Coll et al.[72], and on SNPs described in Shitikov et al. for L2 sublineages[73].

To build the phylogeny, we created a pseudo-alignment with all non-redundant polymorphic positions in the dataset by concatenating all high-quality SNPs, excluding positions associated with drug resistance. SNPs with an allele frequency lower than 90% or positions covered with less than 7 reads were encoded with an “X”. Based on this alignment we built a phylogeny using IQ-TREE 2[74] with the general time-reversible model of sequence evolution, 1,000 bootstrap replicates, indicating the number of invariant sites of each nucleotide.

### Statistical analysis and modeling

The data and a detailed step-by-step description of all analyses performed are available at https://git.scicore.unibas.ch/TBRU/georgia_tb_tx_outcomes. We provide this as supplementary material and refer to it throughout the manuscript. Briefly, to select the variables to be included in the final logistic regression models, we were initially inclusive and considered 66 available clinical and bacterial variables potentially relevant for patient treatment outcomes. Missing data for categorical variables were encoded as “missing”, whereas continuous data for BMI, and TTP was imputed using multivariate imputation by chained equations (MICE). To select for those variables that better explain patient treatment outcome, we performed feature selection based on two methods. First, we conducted a LASSO regularization analysis. Second, we built a random forest model. We included in the initial model the combination of the predictors whose coefficients were not shrinked to zero in the LASSO regularization, and the top ten predictors of the random forest. Model performances were assessed by 10-fold cross validation based on the AUC-PR given the unbalance of the dataset (∼11% of the positive class, unfavorable outcomes). While the AUC-ROC provides limited information in such cases, we report it in this study for comparison with models reported in previous studies.

### GWAS

To find mutations associated with treatment outcomes and disease manifestation, we used the linear mixed effect model of pyseer. The GWAS analyses were adjusted by diagnosis date, patient age, sex, diabetes, hiv-coinfection, employment, alcohol intake, drug abuse, treatment for other diseases and drug resistance profile, as well as by the population structure. To correct for the population structure, we calculated a similarity matrix from the phylogeny using the script “phylogeny_distance.py” provided with pyseer, and specified the sublineage of each MTBC isolate. Additionally, we carried out gene burden tests based on the annotation of *M. tuberculosis* H37Rv. To identify associations correcting by multiple testing, for each GWAS we calculated a p-value threshold by dividing 0.05 over the number of mutations or genes tested, after filtering those that failed chi-square tests or that showed signs of population structure that was not corrected, such as those that showed the exact same values of allele frequency, p-values and estimated coefficients.

## Data Availability

The Supplementary Material is publicly available in a GitLab repository (link below), where we provide supplementary methods, results, the complete dataset, and a detailed step-by-step description of all analyses conducted in R markdown documents.

https://git.scicore.unibas.ch/TBRU/georgia_tb_tx_outcomes

## Acknowledgements

This work was funded by the Swiss National Science Foundation (grants 320030-227432 and CRSII5_213514) and by the European Research Council (grant 883582-ECOEVODRTB).

## Competing interests

The funders of the study had no role in study design, data collection, data analysis, data interpretation, or writing of the report.

## Notes

### Competing Interest Statement

The authors have declared no competing interest.

### Author Declarations

The National Ethics Council under the Ministry of Health of Georgia and the Ethics committee of the National Centre for Tuberculosis and Lung Disease in Tbilisi (Georgia), and the Ethics Commission of North- and Central Switzerland granted ethical approval for this study. The ethics committees waived the need for individual patient consent since only limited and anonymized clinical data were used.

